# Evidence for lower threshold for diagnosis of hypertension: inferences from an urban-slum cohort in India

**DOI:** 10.1101/2021.06.11.21258759

**Authors:** Onkar Awadhiya, Ankit Tiwari, Premlata Solanki, Anuja Lahiri, Neelesh Shrivastava, Ankur Joshi, Abhijit P Pakhare, Rajnish Joshi

## Abstract

**Background:** Hypertension (HTN) is a key risk-factor for cardiovascular diseases (CVDs). Blood-pressure (BP) categorizations between systolic blood pressure (SBP) of 120 and 140 remain debatable. In the current study we aim to evaluate if individuals with a baseline SBP between 130-140 mm Hg (hypertension as per AHA 2017 guidelines) have a significantly higher proportion of incident hypertension on follow-up, as compared to those with SBP between 120-130 mm Hg.

**Methods:** Secondary data analysis was performed in a community-based cohort, instituted, and followed since 2017. Participants were aged ≥30 years, residents of urban slums in Bhopal. BP was measured at or near home by Community Health Workers (CHWs). Two-year follow up was completed in 2019. We excluded participants who were on BP reduction therapy, had fewer than two out-of-office BP measurements and who could not be followed. Eligible participants were re-classified based on baseline BP in four categories: Normal (Category-A), Elevated-BP (Category-B), Variable-BP (Category-C) and reclassified HTN based on AHA-2017 (Category-D). Proportion of individuals who developed incident hypertension on follow up was primary outcome.

**Result:** Out of 2649 records, 768 (28.9%), 647 (24.4%), 586 (22.1%), 648 (24.4%) belonged to Categories A, B, C and D respectively. Incident HTN with cut-off of 140/90 mm Hg was, 1.6%, 2.6%, 6.7%, 12% in categories A, B, C and D respectively. Incidence of incident hypertension in individuals with a baseline SBP between 130-140 mm Hg (Category D) was significantly higher as compared to those with SBP between 120-130 mm Hg (Category B).

**Conclusion:** We conclude that biological basis for AHA-2017 definition of hypertension is relatively robust also for low income and resource-limited settings. Evidence from our longitudinal study will be useful for policy makers for harmonizing national guidelines with AHA-2017.

## Introduction

Cardiovascular disorders (CVD) are the leading cause of morbidity and mortality worldwide.^1^Hypertension is the most powerful, independent, preventable risk factor for death and disability from cardiovascular diseases.^2^ It is also a leading risk factor for all-cause mortality and the largest contributor to global disability-adjusted life years (DALYs).^3^The current prevalence of hypertension (based on the ≥140/90 mmHg threshold) in India is estimated to be 28.9% in both men and women.^4^ Higher a blood-pressure (BP) value greater is cardiovascular risk. Starting at 115/75 mmHg, CVD risk doubles with each increment of 20/10 mmHg throughout the blood pressure range. Risk of CV death increases twofold if BP rises to 135/85, fourfold if BP rises to 155/95, and eightfold at 175/105.^5,6^

While BP measurement technology is simple and is widely available, its diagnostic cut-offs and accuracy of measurement have been a matter of debate.^7,8^ Final results of SPRINT Study have reaffirmed systolic blood pressure of 120mm Hg as a target to treat, ^9^ further 10mm Hg lower than AHA/ACC 2017 guidelines.^10^ Lowering of diagnostic threshold would increase prevalence of hypertension, and will pose a higher burden on provision of pharmacotherapy. ^11,12^ About a third of SPRINT study participants had their Systolic BP below 132mm hg, between 132-145mm Hg and above 145mm Hg respectively, and those in the lowest tertile had greatest benefit by intensive treatment [HR 0.70 (95%CI 0.51-0.95)].^13^ In SPRINT study, BP was measured by automated oscillometric measurement, which minimizes the effect of white coat hypertension, unlike blood-pressure measured in most clinical practice settings.^14^

While SPRINT demonstrated lowering of BP control threshold, and its consequent pharmacotherapy to be beneficial, it was achieved in a setting of more accurate BP-measurement. This study prompted AHA/ACC in 2017 to lower the diagnostic threshold from previous 140/90 to 130/80mm Hg.^10^ In 2018, European guidelines revised its guidelines, and elaborated on thresholds based on measurement technique.^15^ These guidelines have caused uncertainty in diagnosis of hypertension. An individual with a systolic BP (SBP) between 120 and 140 mm Hg or diastolic BP (DBP between 80 and 89mm Hg) may have either true hypertension or a white-coat hypertension or no-hypertension, depending on measurement technique or guideline applied for decision-making.^16^It has been demonstrated that as many as 70% of all white-coat hypertension, and 95% of masked hypertension falls between this systolic blood pressure-range.^17^Arguments for clinical benefit of a lower blood-pressure threshold are early recognition of disease and mortality benefit, with an implicit assumption that if untreated, BP of these individuals will continue to rise.^18^

National guidelines in India have not yet adopted AHA 2017 guidelines for hypertension, and it has been debated that addition of more individuals with newer definitions will overwhelm our Public health delivery system.^19,20^ Lack of longitudinal studies and potential misclassification in blood-pressure values at lower diagnostic cut-offs were identified as concerns.^21^

We had established a population based cohort in 2017, to evaluate CVD risk-factors and their control.^22^ Community health workers (CHWs) obtained multiple sets of BP measurements and all therapeutic decisions were based on diagnostic threshold of SBP >/= 140 or DBP >/= 90 at baseline, which remains the current national guideline. If AHA/ACC 2017 guidelines are applied, individuals with BP below this threshold would get re-classified as having elevated blood-pressure (SBP between 120 and 130mm Hg) or hypertension (SBP between 130 and 140mm Hg). We completed a two-year follow up of cohort in 2019,^23^ and a secondary data analysis will enable us to evaluate blood-pressure outcomes in re-classified cohort members. Our key research question is, among cohort members with at least two out-of-office BP assessments at baseline, do individuals with SBP values between 130 and 140, (as compared to those with SBP between 120 and 130mm Hg) have a significantly higher proportion of incident hypertension or mean SBP at the end of two years. Our hypothesis is that a robust classification system will exhibit a consistent trend in blood-pressure outcomes on follow up.

## Methods

### Design

The current study is a secondary data analysis from a community based cohort that was established in year 2017, and is being followed up thereafter.^23^ Primary study design was approved by institutional ethics committee, and participants who provided a written informed consent were included in the study.

### Setting

Cohort-members include consenting non-pregnant adults, aged 30 years or more, residing in 16-different urban slums in city of Bhopal, India. At baseline in year 2017-18, eligible participants had their CVD-risk assessment done based on interview, anthropometry, and BP measurement. BP was measured on multiple occasions in all participants, which consisted of an average of three values obtained one minute apart, on each occasion. First set of blood-pressures were measured by CHWs at home of all the participants, second set was obtained by study supervisors in neighbourhood camps and a third set was obtained in public health facilities for confirmation of hypertension status. First two of these BPs were out-of-office measurements. We completed a two-year follow up of all participants in 2019, and obtained one-set of home BP measurement for all participants and a confirmatory home BP measurement for those with newly detected hypertension above 140/90 mm Hg. All BP measurements were obtained using a digital sphygmomanometer (Omron digital apparatus model 7200, Kyoto, Japan).

### Data source

The data of the primary study was collected using a Commcare based mobile application, and is stored in cloud based secure servers. A dataset, stripped of all personal identifiers and consisting of required variables was extracted for the current study. This data-set contains information on a total of 6174 individuals, and includes variables pertaining to demography (age, gender, education, wealth index classification), blood-pressure values obtained at different time-points, and CVD-risk assessment at baseline (tobacco use, physical activity status, prior CVD history, waist circumference, body mass index). The data-set also contains information about initiation of anti-hypertensive pharmacotherapy, linkage to facilities, adherence and monitoring of those on therapy.

### Procedures and definitions

We identified those individuals who had their BP measured at-least twice at baseline in 2017. It was required that BP on both occasions at baseline was obtained in out-of-office setting, in order to be comparable to follow-up measurements obtained in 2019, which were all home based. Hence, Individuals who had one out-of-office and one facility level BP measurement at baseline, and those who did not have a follow up value were excluded. We also excluded individuals who were advised any pharmacological or non-pharmacological measures for blood-pressure reduction. Thus, individuals who had two BP values above therapeutic threshold of 140/90 mm Hg at baseline or those on a BP-lowering medication for alternate indication (such as for ischemic heart disease, ischemic stroke, diabetes nephropathy) were excluded. Thus, data-set for the current study consisted of adults above age of 30-years, who were therapy naïve, and had multiple comparable out-of-office BP values at baseline and on follow-up.

We used baseline out-of-office BP values and re-classified individuals into four different blood-pressure categories, Normal (Category A), Elevated blood pressure based on AHA 2017 (Category B), Variable blood pressure (Category C) and reclassified hypertension based on AHA 2017 (Category D). (Table 1) Our primary outcome was proportion of incident hypertension (SBP >/= 140mm Hg or DBP >/= 90mm Hg). We used this definition, as all individuals in our data-set had their BPs below this threshold at baseline. Hypertension as per AHA 2017 definition (SBP >/= 130mm Hg or DBP >/= 80mm Hg) and mean SBP were our secondary outcomes.

**Table 1:** Blood-pressure categories and their definitions in the current study.

| Category | Name | Definition |  |
| --- | --- | --- | --- |
|  |  | Systolic Blood pressure | Diastolic Blood Pressure |
| A | Normal | Less than 120mm Hg on two occasions | Less than 80mm Hg on two occasions |
| B | Elevated BP based on AHA/ACC definition* | 120-129 mm Hg on two occasions, or once in this range and once below 120mm Hg | Less than 80mm Hg on two occasions |
| C | Variable BP | Above 130 mm Hg on one occasion, and between 120-129mm Hg on another | Between 80-89mm Hg on one occasion, or less than 80mm Hg on two occasions |
| D | Reclassified Hypertension based on AHA/ACC definition* | 130-139mm Hg on two occasions, or once in this range and once 140mm Hg or more. | 80-89 mm Hg on two occasions, or once in this range and once 90mm Hg or more. |

\* AHA/ACC definition as per reference (10) BP=Blood pressure

### Statistical analysis

Our primary outcome was proportion of individuals who developed incident hypertension on follow up. Our key comparison was between categories B, C and D, with category A as a reference. We also performed a descriptive analysis, comparing CVD-risk factors amongst individuals in each category, and used appropriate tools for data visualization. We used chi-square test to compare nominal variables, One way ANOVA or Kruskal Wallis test followed by post-hoc pair wise comparisons for numerical variables. p-value of less than 0.05was considered as statistical significant. Analyses were conducted using the R Statistical language (version 4.0.3; R Core Team, 2020) on macOS Catalina 10.15.6, using the packages gtsummary (version 1.4.1; Daniel Sjoberg et al., 2021),^24^ggforce (version 0.3.2; Thomas Lin Pedersen, 2020)^25^ and tidyverse (version 1.3.0; Wickham et al., 2019).^26^

## Results

Of a total of 2649 records who met the study criteria, 648 (24.4%) individuals had BP above the AHA 2017 defined threshold of 130/80mm Hg at baseline, and were labelled as ‘re-classified HTN’(category D). Another 647 (24.4%) individuals had ‘elevated BP’ (category B), and 586 (22.1%) had a ‘variable’ BP between these two groups (category C). Remaining 768 (28.9%) individuals had a normal BP (Category A). Individuals in highest BP-category (Category D) were older (median age 42 years), had a higher proportion of men (47%), had more tobacco-users (38%), higher prevalence of dysglycemia (23%) or abdominal obesity (62%). These CVD-risk factors (higher age, male-gender, tobacco-use, dysglycemia, obesity) demonstrated a rising trend across BP-categories. (Table 2)

**Table 2:** Baseline and characteristics of the study population.

| Characteristic | Overall, N =<br>2,649<sup>1</sup> | Normal<br>(SBP<120),<br>N = 768<sup>1</sup> | Elevated<br>(SBP 120-129),<br>N = 647<sup>1</sup> | Variable,<br>N = 586<sup>1</sup> | Reclassified HTN<br>(SBP 130-139)<br>N = 648<sup>1</sup> | p-value |
| --- | --- | --- | --- | --- | --- | --- |
| BP-category |  | A | B | C | D |  |
| Median Age | 40 (35, 48) | 36 (32, 44) | 40 (34, 47) | 42 (35, 50) | 42 (36, 50) | <0.001 |
| Male Gender | 957 (36%) | 180 (23%) | 247 (38%) | 228 (39%) | 302 (47%) | <0.001 |
| Wealth Tertile |  |  |  |  |  |  |
| T1 | 704 (28%) | 215 (29%) | 155 (25%) | 153 (27%) | 181 (29%) |  |
| T2 | 823 (32%) | 261 (36%) | 213 (34%) | 171 (30%) | 178 (29%) | 0.009 |
| T3 | 1,015 (40%) | 254 (35%) | 254 (41%) | 245 (43%) | 262 (42%) |  |
| Tobacco use | 868 (33%) | 228 (30%) | 201 (31%) | 196 (33%) | 243 (38%) | 0.012 |
| Dysglycemia | 451 (17%) | 89 (12%) | 113 (17%) | 103 (18%) | 146 (23%) | <0.001 |
| Abdominal<br>Obesity | 1,351 (51%) | 294 (38%) | 339 (52%) | 318 (54%) | 400 (62%) | <0.001 |
| Median Waist<br>circumference | 84 (75, 91) | 78 (70, 85) | 84 (77, 92) | 84 (76, 92) | 87 (80, 95) | <0.001 |
| Median Body<br>Mass Index | 23.3 (20.3, 26.2) | 21.5 (19.0,<br>24.6) | 23.6 (20.8,<br>26.5) | 23.5 (20.9,<br>26.4) | 24.4 (21.9, 27.7) | <0.001 |

Outcomes were also significantly different across BP-categories. Proportion of individuals who developed incident hypertension with a diagnostic threshold of 140/90 mm Hg was 1.6%, 2.6%, 6.7%, 12%and in categories A, B, C and D respectively. Proportion of individuals with incident hypertension were higher in each-category with a lower diagnostic threshold of 130/80 mm Hg. The trend of higher incident-hypertension across BP-categories persisted with lower diagnostic threshold. (Table 3)

**Table 3:** Outcome measures in study population.

| Characteristic | Normal<br>(SBP<120),<br>N = 768<sup>1</sup> | Elevated<br>(SBP 120-129),<br>N = 647<sup>1</sup> | Variable,<br>N = 586<sup>1</sup> | Reclassified<br>HTN<br>(SBP 130-139)<br>N = 648<sup>1</sup> | p-value<sup>2</sup> |
| --- | --- | --- | --- | --- | --- |
| BP-category | A | B | C | D |  |
| Mean Baseline SBP (SD) | 107.8 (7.4) | 118.9 (6.9) | 123.2 (12.2) | 131.5 (10.1) | <0.001 |
| Mean Follow-up SBP (SD) | 114.4 (11.7) | 122.7 (11.9) | 128.1 (14.8) | 133.5 (14.8) | <0.001 |
| Incident Hypertension<br>(>130/80 mm Hg) | 22 (3.5%) | 38 (9.5%) | 71 (21%) | 130 (44%) | <0.001 |
| Incident Hypertension<br>(>140/90 mm Hg) | 12 (1.6%) | 17 (2.6%) | 39 (6.7%) | 76 (12%) | <0.001 |

*p-value for A\*B for Incident Hypertension 0.159, for rest all pairwise comparisons p<0.001*

Over a two-year period BP-categorization was unchanged in 1068 (40.3%) individuals. Those in categories A and D (normal and re-classified HTN) were most stable as 68% and 62% of them were in the same BP-category on follow-up. BP-categorization was dynamic in categories B and C (elevated and variable BP) as only 145 (11.8%) individuals were in the same BP-category on follow-up. (Figure 1)

**Figure 1:**
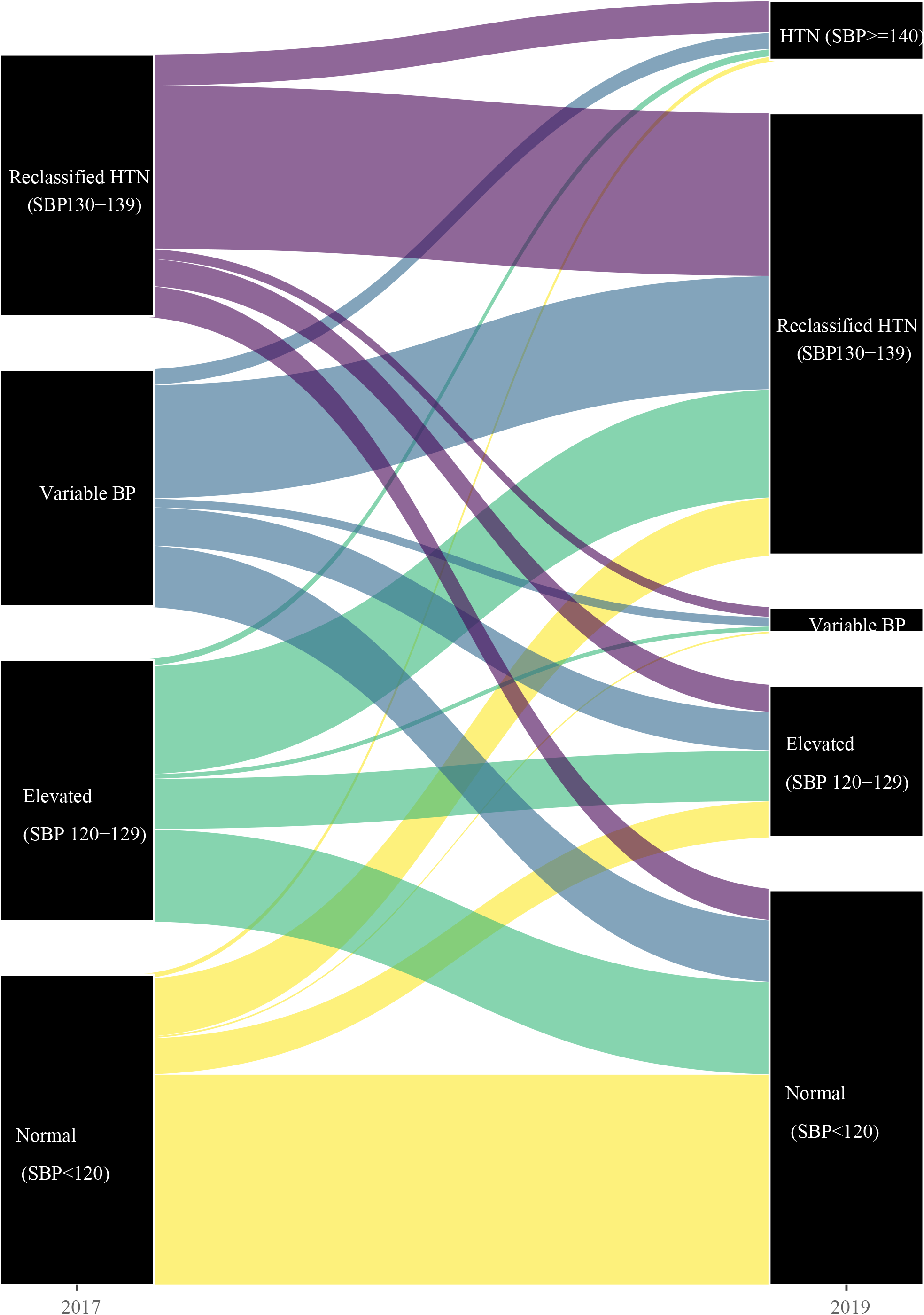
Trajectory of BP classification from 2017 to 2019.

## Discussion

We found that BP-categorization as per AHA 2017 is robust, as individuals re-classified as having HTN based on diagnostic threshold of 130/80mm Hg not only have a higher prevalence of CVD-risk factors at baseline, are also more likely to develop incident HTN. Previously entire 20mm Hg spectrum of SBP between 120-139 was categorized as ‘pre-hypertension’ which was subdivided into two 10mm Hg bands of elevated-BP (SBP between 120-129) and stage I hypertension (SBP between 130-139) as per AHA 2017 classification. We found that difference in outcomes in these 10mmHg bands are large, as only 2.6% Individuals with ‘elevated-BP’ developed HTN (above 140/90mm Hg) as compared to 12% in those with re-classified stage I hypertension.

In our study BP-levels were measured by CHWs using digital sphygmomanometers. Accuracy in BP measurement has been a concern when we attempt to classify individuals in narrow 10mm Hg bands. It is known that office-BP measurement has its inaccuracies,^8^ and out-of-office BP is better as it would reduce white-coat hypertension.^27^However inter-instrument variability has been a concern for home-BP measurements.^28^ We used an intermediate strategy, wherein BP is measured using same instrument by community-health workers (CHWs) at or near the home of the participants.While accuracy of such measurements in comparison to 24-hour ambulatory blood pressure measurement (ABPM) is not established, observed differences in our study even in narrow 10mm Hg bands validates these measurements.

We encountered BP-variability in about one-fifth of our participants at baseline. These were individuals who had their SBP between 120-129 mm Hg on one occasion, and between 130-139 mm Hg on another. One explanation of this variability is location of BP-measurements as one was a home-based measurement and another was obtained in the neighborhood. Proportion of individuals with a variable BP was low on follow-up as all measurements were obtained at home.(figure 1) BP is a dynamic physiological parameter which is affected by various factors like volume status, cardiac functions, emotional state, sleep-awake status, time of the day, season or the age of the person. ^29^High BP variability has gained attention as an independent CVD risk factor.^30^While we currently believe that two BP-values need to be concordant for a more robust diagnosis of hypertension, diagnostically uncertain variable individuals however are a special phenotype.^31^In our study distribution of other CVD risk-factors as well as progression to incident HTN in individuals with a variable BP was intermediate to those with ‘elevated-BP’ or those with ‘re-classified HTN’. This finding also strengthens the concept that long-term variability is a biological characteristic rather than a measurement error. It however remains debatable if benefits of anti-hypertensive treatment should be extended to this group as well. ^32^

Purpose of revision of diagnostic threshold has been to identify a minimum level which is associated with adverse impact on the bodily functions, target organ damage and increased mortality and at which any intervention to treat is likely to be associated with reduction of its ill effects.In 2017 AHA released new guidelines for the prevention, detection, evaluation and management of hypertension which used systolic BP of 130 mmHg or higher and diastolic BP of 80 mmHg or higher to classify individuals as hypertensive. While JNC-7 guidelines which were in common use since 2003 used systolic BP of 140 mmHg or higher and diastolic BP of 90 mmHg or higher for the same.Measurement of blood pressure in entire communities helps determine burden of disease, which is expected to become higher with reduced diagnostic thresholds. In present study, about a quarter of individuals who were previously classified as ‘not having hypertension’ as per JNC-guidelines, satisfied revised AHA 2017 criteria to be re-classified as hypertension. In a previous study, with a diagnostic threshold of 140/90 mm Hg, we estimated that 25% of adults have hypertension.^33^This proportion would become about 44% with a lower threshold of 130/80 mm Hg. This increment is similar as in other settings, and lowering of threshold also increases the absolute number of individuals with blood-pressure unawareness, and those with white-coat hypertension.^34–36^This would increase the burden on health-systems, and push for lowering of BP-control closer to therapeutic threshold of 120/80 mm Hg. (Figure 2) While, this is challenging but need to harmonize various hypertension guidelines to a diagnostic and therapeutic threshold of 130/80mm Hg is unavoidable. ^37^

**Figure 2:**
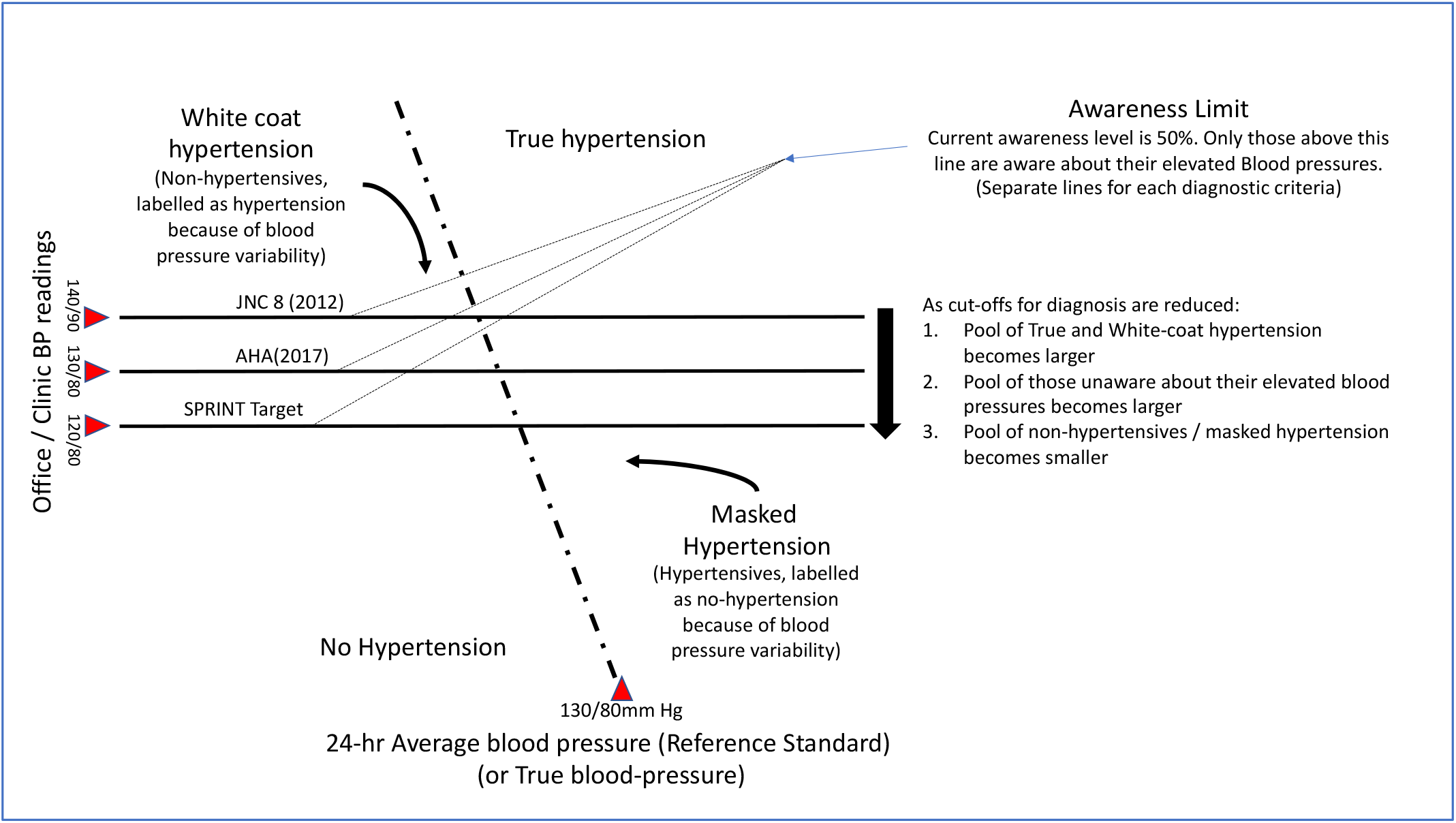
Blood-pressure categories and hypertension classification thresholds.

Our study has several limitations. First, only two out-of-office BP measurements were available, and presence of a third similarly obtained value would have reduced the proportion of those with a variable BP. Thus, we might have overestimated the proportions of individuals with a ‘variable BP’ at baseline. Second, a complete set of BP values was available in only about two-fifths of the entire cohort. Thus, our inferences are generalizable only to population sub-groups that are adherent to follow-ups and repeat BP measurements. Third, rise in blood-pressure is only an intermediate outcome and our study was not designed to evaluate hard-CVD outcomes such as target organ damage or mortality. Despite these limitations, a key strength of the current study is using available cohort data from a low-income setting, where BP was measured with minimal sophistication, to demonstrate robustness of BP-categorizations below the diagnostic threshold of 140/90mm Hg.

To conclude, biological basis for AHA 2017 definition of hypertension is reasonably strong, even in low-income settings where BP measurements are done with minimal resources. Individuals with elevated and variable BP are also distinguishable biological categories. The results of our study can be generalised to resource constrained conditions such as ours. Hence, use of a simple digital sphygmomanometer and application of lower threshold of BP cut-offs can be a more practical way to identify individuals with hypertension unawareness, higher likelihood of incident hypertension and individuals with variable BP. With demonstration of mortality-benefit of treating individuals with lower SBP as demonstrated in SPRINT study, there is a need to apply these concepts for better patient-care. Evidence from our longitudinal study will be useful for policy makers for harmonizing national guidelines with AHA 2017.

## Supporting information

STROBE checklist

## Data Availability

Raw data of this study is not deposited in any public repository. However, anonymized raw data of this study would be available to academicians or researchers on reasonable request to corresponding author.

## Funding

This study was funded by Indian Council of Medical Research, New Delhi as an extramural project grant. Funders have no role in data collection, analysis and writing of the manuscript. (Grant – PI-Dr Rajnish Joshi, IRIS-2014-0976)

## Ethics approval

The study design was approved by the Institutional Human Ethics Committee of All India Institute of Medical Sciences, Bhopal (Ref: IHEC-LOP/2017/EF00045).

## Informed consent

Participant Information Sheet in Hindi language was provided to each participant. All participants provided written informed consent prior to initiation of any study procedures.

## Author contributions

RJ conceived the study; APP, AT, PS and OA developed the protocol; ALand NS acquired data, AJ, APP and RJ supervised data acquisition, APP, OA and RJ analyzed data and wrote first draft. All authors critically reviewed the first draft and provided inputs for its revisions.

## Notes

### Competing Interest Statement

The authors have declared no competing interest.

